# Clinical Trial Data Sharing: A Cross-Sectional Study of Outcomes Associated with Two NIH Models

**DOI:** 10.1101/2021.09.10.21263404

**Authors:** Anisa Rowhani-Farid, Alexander C. Egilman, Audrey D. Zhang, Cary P. Gross, Harlan M. Krumholz, Joseph S. Ross

**Affiliations:** Restoring Invisible and Abandoned Trials Support Center, Department of Pharmaceutical Health Services Research, University of Maryland, Baltimore, 220 N Arch St., Baltimore MD, 21201, United States of America (USA); Program On Regulation, Therapeutics And Law (PORTAL), Division of Pharmacoepidemiology and Pharmacoeconomics, Brigham & Women’s Hospital, Harvard Medical School, 1620 Tremont Street, Suite 3030, Boston, MA 02120, USA; Department of Internal Medicine, Duke University School of Medicine, 2301 Erwin Rd., Durham, NC 27710, USA; Section of General Internal Medicine, Yale School of Medicine, 333 Cedar St., New Haven, CT 06510, USA; Cancer Outcomes Public Policy and Effectiveness Research (COPPER) Center, Yale School of Medicine, 367 Cedar St., New Haven, CT 06520, USA; National Clinician Scholars Program, Yale School of Medicine, 333 Cedar St., New Haven, CT 06510, USA; Center for Outcomes Research and Evaluation (CORE), Yale-New Haven Hospital, 1 Church St., Suite 200, New Haven, CT 06510, USA; Section of Cardiovascular Medicine, Yale School of Medicine, 333 Cedar St., New Haven, CT 06510, USA; Department of Health Policy and Management, Yale School of Public Health, 60 College St., New Haven, CT 06520, USA

## Abstract

**Background:** The impact and value of clinical trial data sharing, including the number and quality of publications that result from shared data – “shared data publications” – may differ depending on the data sharing model used.

**Methods:** We characterized the outcomes associated with two data sharing models previously used by Institutes of the U.S. National Institutes of Health (NIH): NHLBI’s centralized model, which uses a repository to manage data sharing requests, and NCI’s decentralized model, which entrusted research groups to independently manage data sharing requests. We identified trials completed in 2010 that met NIH data sharing criteria and matched studies sponsored by each Institute based on cost or size, determining whether trial data were shared and the frequency of shared data publications.

**Results:** We identified 14 NHLBI-funded trials and 48 NCI-funded trials that met NIH data sharing criteria. We matched 14 NCI-funded trials to the 14 NHLBI-funded trials; among these, 4 NHLBI-sponsored trials (29%) and 2 NCI-sponsored trials (14%) shared data. From the 2 NCI-sponsored trials sharing data, we identified 2 shared data publications, one per trial, both of which were meta-analyses. From the 4 NHLBI-sponsored trials sharing data, we identified 7 shared data publications, all using data from 1 trial, 5 of which were pooled analyses and 2 reported secondary outcomes.

**Conclusion:** When characterizing the outcomes associated with two NIH data sharing models, both the NHLBI and the NCI models resulted in only 21% of trials sharing data and few shared data publications. There are opportunities to optimize clinical trial data sharing efforts both to enhance clinical trial data sharing and increase the number of shared data publications.

## Introduction

Clinical trial data sharing involves sharing data generated from a clinical trial, which includes protocols, informed consent forms, case report forms, clinical study reports, and individual patient-level data (IPD). Sharing clinical trial data has numerous benefits, including creating opportunities to pursue additional research questions, meta-analyses, and independent verification and reproducibility of study results.^1–3^ Data sharing also aims to maximize the value of clinical trial data by strengthening the quality and totality of the evidence driving medical decisions.^4^ In recent years, sharing clinical trial data, particularly IPD, has become more commonplace among scientific researchers and sponsors, catalyzing the development of various models for sharing IPD to researchers independent of original investigational teams.^5–9^

IPD data sharing may occur through one of several frameworks.^8^ In a centralized model, IPD are prepared and released into a central repository with access overseen by an independent entity, commonly a private or public funder, foundation, or academic institution, that reviews and approves data requests; IPD is then sent to investigators.^10^ Alternatively, in a decentralized model, investigators independently or as part of a research collaborative retain control of their data and grant access to data requests at their discretion. A third approach involves unrestricted open access, where IPD are made freely available to independent researchers without a gatekeeper. Due to patient privacy issues, it is not common for public repositories with unrestricted open access to store IPD from clinical studies. As such, clinical trial data are usually shared via a decentralized or centralized model.

The NIH has been a long-standing proponent of data sharing, instituting an initial data sharing policy in 2003 requiring investigators to submit detailed data sharing plans as a condition for funding clinical trials, regardless of size, requesting $500,000 or more in direct costs in any year of the proposed project, and finalizing a data sharing policy in 2020 (effective 2023) requiring all NIH-funded research to submit a Data Management and Sharing Plan.^11,12^ While NIH strongly encouraged IPD sharing as part of these policies, it was not required. Instead, individual NIH institutes developed their own models for data sharing^13^ and the NIH has yet to mandate data sharing for clinical trials across the Institutes,^12,14^ despite calls by leaders in the field for NIH to take on the role of leading the data sharing movement, with an initial focus on clinical trials.^15^

Characterizing the outcomes of differing data sharing models and their impact may help inform future data sharing model structures and policies^16^. The NHLBI and NCI, as two examples, differ in their approaches to data sharing despite both being part of the NIH. For example, the NHLBI has had a data-sharing policy in place since 1989 that ultimately resulted in the creation of the NHLBI Data Repository in 2000, a centralized model for housing clinical data from NHBLI-sponsored studies,^11^ managed under the NHLBI Biologic Specimen and Data Repository Information Coordinating Center (BioLINCC) since 2008.^17,18^ Through BioLINCC, NHLBI-sponsored investigators with 500 or more participants in their studies or those requesting $500,000 or more in direct costs in any year of the proposed project must deposit IPD into a central repository; all data sharing requests are reviewed by NHLBI staff and are evaluated based on the inclusion of a description of the research plan/protocol, as well as documentation of review or or an exemption from review from an Institutional Review Board or Ethics Committee.^11^ In contrast, up until February 2017, NCI had adopted a more de-centralized approach to data sharing, relegating control to large cooperative groups responsible for managing the data collected through their own studies. Each cooperative group maintained an electronic database housed at the Group’s Statistical Center that aggregated data from participating institutions.^19^ Outside investigators could request IPD by submitting a formal request to the Group, which then could grant approval after internally reviewing the “scientific merits and feasibility” of the research proposal. Requests were considered only for data for which the primary study analyses had already been published.^19^

While the NCI data sharing model has since been updated to more closely approximate NHLBI’s,^20^ the initial use of two different models by these NIH institutes allows for their comparison to determine their relative outcomes.^21^ The true value in sharing data lies in the generation and dissemination of new knowledge resulting from shared data, potentially best represented by “shared data publications”, peer-reviewed research studies based on independent analyses of shared IPD and authored by investigators external to the primary study team. Accordingly, we characterized the outcomes associated with the NHLBI and NCI models of data sharing, determing the number of shared data publications and other metrics of research value for a matched sample of completed trials sponsored by each institute that were completed in 2010. Evaluating the “outcomes” of existing initiatives via examination of shared data publications will aid in understanding how and to what extent IPD are being shared and used.

## Results

We identified two trials with a primary completion date between January 1, 2010 and December 31, 2010 with total costs greater than or equal to $1,000,000 that had enrolled 500 or more patients. Next, we identified a total of 53 clinical studies (50 NCI-sponsored and 3 NHLBI-sponsored) with a primary completion date between January 1, 2010 and December 31, 2010 with total costs greater than or equal to $1,000,000, although none had enrolled 500 or more patients. Of these, according to ClinicalTrials.gov, 2 NCI-sponsored trials were withdrawn, 13 were terminated, and 1 was no longer available, whereas 1 NHLBI-sponsored trial was terminated; the remaining trials listed their status as completed. We then identified 11 NCI-sponsored trials and 13 NHLBI-sponsored trials that had 500 or more participants. Of these, 3 NCI-funded trials and 2 NHLBI-funded trials had no cost information available, the remaining trials had total costs less than $1,000,000 in any given fiscal year. Out of these trials with 500 or more participants, 3 NCI-funded trials and 1 NHLBI-funded trial had an unknown status; the remaining trials listed their status as completed.

In total, we identified 42 NCI-sponsored trials and 14 NHLBI-sponsored trials with a primary completion date between January 1, 2010 and December 31, 2010 that listed their status as completed and either had total costs greater than or equal to $1,000,000 or had enrolled 500 or more patients. We matched the 14 NHLBI-funded trials to 14 NCI-funded trials that had similar completion dates, sample size and costs; see flow chart depicted in Figure 1. Among the 14 NHLBI-sponsored trials, 6 were interventional and 8 observational, whereas among the 14 NCI-sponsored trials, 7 were interventional and 7 observational; Table 1 summarizes the characteristics of the 28 trials, stratified by funder.

**Table 1:** Characteristics of the matched trials, stratified by funder

|  | Funder |  |  |  |
| --- | --- | --- | --- | --- |
|  | NHLBI (n = 14) |  | NCI (n = 14) |  |
| **Trial Characteristics** | *n* | % | *n* | % |
| **Sample size** |  |  |  |  |
| 1-1000 | 9 | 64 | 12 | 86 |
| 1001-2000 | 3 | 21 | 1 | 7 |
| 2001-3000 | 1 | 7 | 0 | 0 |
| > 3001 | 1 | 7 | 1 | 7 |
| **Total Costs** |  |  |  |  |
| < 1,000,000 | 9 | 64 | 4 | 29 |
| ≥ 1,000,000 | 3 | 21 | 7 | 50 |
| Cost data not available | 2 | 14 | 3 | 21 |
| **Study type** |  |  |  |  |
| Observational | 8 | 57 | 7 | 50 |
| Interventional | 6 | 43 | 7 | 50 |

**Figure 1:**
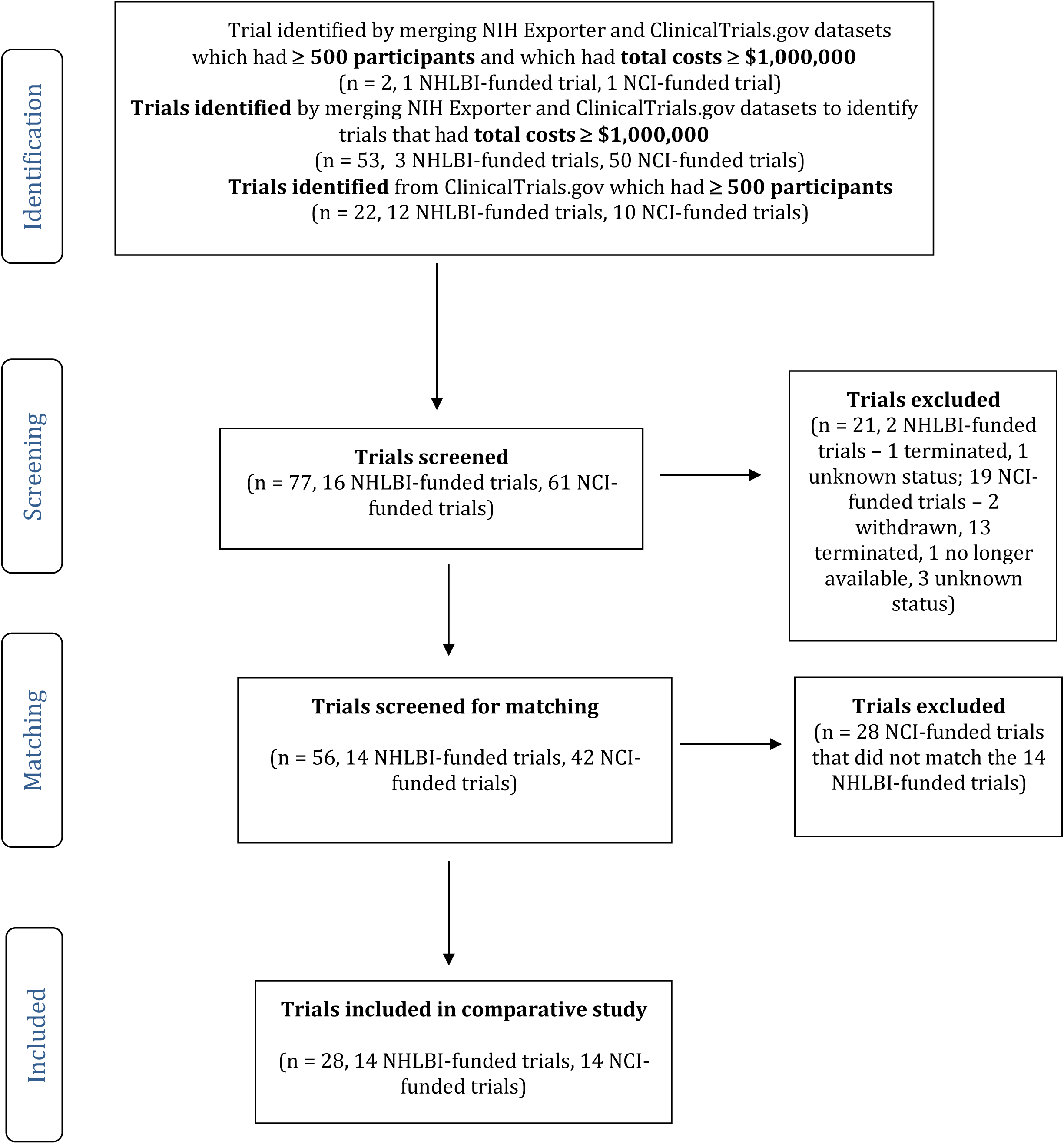
Flow chart demonstrating how the final sample, 14 NHLBI-funded trials and 14 NCI-funded trials, included in the comparative study was created

### Results reporting, publication, and sharing of individual patient-level data

#### NHLBI-funded trials

Among the 14 NHLBI-funded trials, 3 (21%) reported their primary results on ClinicalTrials.gov, and 13 (93%) had their primary results published. The median time from primary study completion to result dissemination through a primary publication was 2 years (IQR, 1.5-3.9 years). In total, 4 of 14 (29%) NHLBI-funded trials shared their data. IPD were available for request on the BioLINCC website for 3 of 14 trials, and at the database for Genotypes and Phenotypes (dbGap) for 1 trial. 5 of 14 authors clarified in their responses that data were not shared and we did not hear back from 5 of 14 authors (2 emails bounced back).

#### NCI-funded trials

Among the 14 NCI-funded trials, 4 (29%) reported their primary results on ClinicalTrials.gov, and 11 (79%) had their primary results published. The median time from primary study completion to result dissemination though a primary publication was 1.6 years (IQR, 0.5-2.8 years). None of the NCI trials shared their data at the National Clinical Trials Network/NCI’s Community Oncology Research Program (NCTN/NCORP) Data Archive. Personal communication with authors of these NCI-funded studies confirmed that 2 of 14 (14%) trials had shared their data with other authors/institutions who were conducting meta-analyses requiring raw data. 5 of 14 authors clarified in their responses that data were not shared and we did not hear back from 7 of 14 authors (one email bounced back).

### Shared data publications

We determined that 4 of 14 (29%) NHLBI-funded trials had secondary result publications generated from internal authors, with a median of 0 (IQR, 0-3.7) internal secondary publications. Only 1 NHLBI-funded trial had any associated shared data publications, the *Diuretic strategies in patients with acute decompensated heart failure* (DOSE-AHF) trial. Similarly, we determined that 6 of 14 (43%) NCI-funded trials had secondary result publications generated from internal authors, with a median of 0 (IQR, 0-2) internal secondary publications. 2 NCI-funded trials had associated shared data publications (both of which were meta-analyses); these two trials were the *Disease Management for Smokers in Rural Primary Care* trial and the *Enhancing Tobacco Use Treatment for African American Light Smokers trial*. Table 2 reports these key outcomes, stratified by funder.

**Table 2:** Results reporting, data sharing, and publication practices of trials funded by NHLBI and NCI

| Key Outcomes | Funder |  |
| --- | --- | --- |
|  | NHLBI (n=14) | NCI (n=14) |
| Proportion (%) reporting primary results on ClinicalTrials.gov | 3 (21%) | 4 (29%) |
| Proportion (%) publishing primary results | 13 (93%) | 11 (79%) |
| Median (IQR) time (years) from study completion to disseminating results through publication | 2 (1.5-3.9) | 1.6 (0.5-2.8) |
| Proportion (%) sharing data | 4 (29%) | 2 (14%) |
| Proportion (%) with secondary result publications (internal team) | 4 (29%) | 6 (43%) |
| Proportion (%) with shared data publications (external team) | 1 (7%) | 2 (14%) |
| Median (IQR) number of secondary result publications (internal team) | 0 (0-3.7) | 0 (0-2) |
| Median (IQR) number of shared data publications (external team) | 0 (0-0) | 0 (0-0) |

We conducted a deep dive into the 3 trials for which we identified multiple shared data publications.

The DOSE-AHF trial had 24 total secondary publications, 17 of which were conducted by internal authors and 7 by external authors, representing shared data publications (Table 3). Of the 17 secondary publications published by internal authors from the DOSE-AHF trial, 13 were pooled analyses, 2 reported secondary outcomes, 1 was a sub-group analysis and 1 a meta-analysis. These secondary internal publications had a median citation count of 10 (IQR, 7-22); median impact factor of 4.3 (IQR, 3.9-6.0); median abstract views of 24 (IQR, 5-67); and median Altmetric Attention Score of 4 (IQR, 0-7). Of the 7 shared data publications, 5 were pooled analyses and 2 reported secondary outcomes. These shared data publications had a median citation count of 17 (IQR, 9.5-37); median impact factor of 3.9, (IQR, 3.9-5.0); median abstract views of 102, (IQR, 70-214.5); and median Altmetric Attention Score of 3 (IQR, 0-5) (Table 3). Figure 2 is a timeline of the secondary publications from the DOSE-AHF trial and the impact factor of the publishing journals, plotted on a logarithmic scale. The only outlier in terms of impact factor was the original publication in the *New England Journal of Medicine*, which has an impact factor of 74.7.

**Table 3:** Types of analyses and impact measures of secondary publications authored by internal and external researchers for the trial: Diuretic strategies in patients with acute decompensated heart failure (NHLBI-funded)

| Types of analyses and impact measures | Authorship |  |
| --- | --- | --- |
|  | Internal<br>( n = 17 publications) | External<br>(n = 7 publications) |
| Secondary analysis type | 13 pooled analyses; 2 secondary outcome reporting; 1 sub-group analysis; 1 meta-analysis | 5 pooled analyses; 2 secondary outcome reporting |
| Median (IQR) citation count of secondary result publications | 10 (7-22) | 17 (9.5-37) |
| Median (IQR) impact factor of publishing journal of secondary result publication | 4.2 (3.9-6.0) | 3.6 (3.6-4.5) |
| Median (IQR) abstract views of secondary result publication | 24 (5-67) | 102 (70-214.5) |
| Median (IQR) Altmetric Attention Score of secondary result publication | 4 (0-7) | 3 (0-5) |

**Figure 2:**
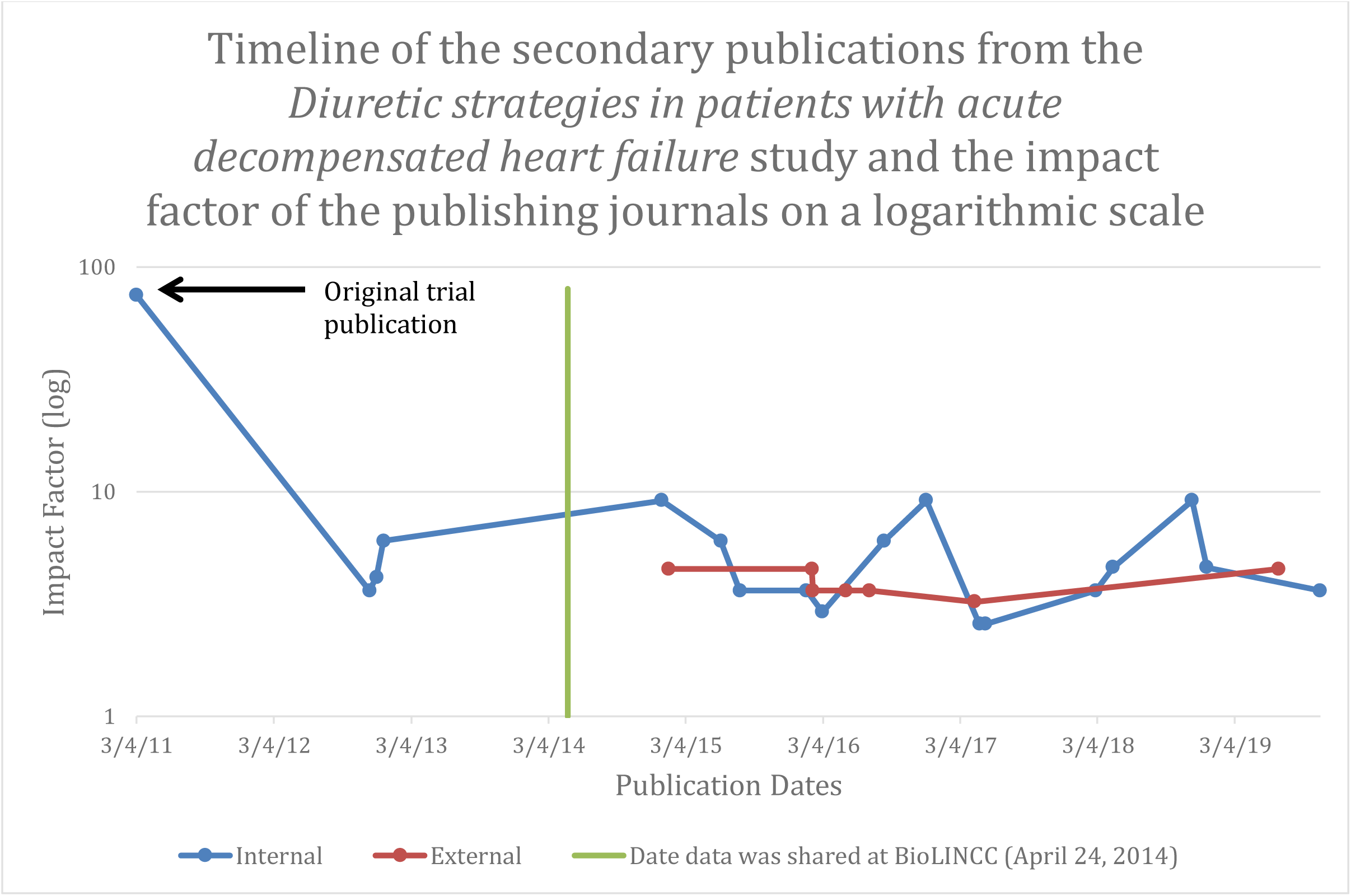
Timeline of the secondary publications from the Diuretic strategies in patients with acute decompensated heart failure study and the impact factor of the publishing journals on a logarithmic scale Notes: The data from this trial was shared at BioLINCC on April 24, 2014 (green vertical line). The impact factor of the publishing journals for the shared data publications that were authored by internal vs. external researchers were comparable (blue and red lines, respectively).

The NCI-funded trial, *Disease Management for Smokers in Rural Primary Care*, had 3 total secondary publications, 2 of which were conducted by internal authors (an extended follow-up study and a modeling study) and 1 by external authors (a meta-analysis) (Table 4). The two internal secondary publications had a median citation count of 4 (IQR, 2-6); median impact factor of 2.8 (IQR, 2.8-2.9); median abstract views of 1091 (IQR, 575-1607); and median Altmetric Attention Score of 0.5 (IQR, 0.25-0.75). The meta-analysis had a citation count of 47, was published in a journal with an Impact Factor of 4.4, had 798 abstract views, and an Altmetric Attention Score of 8. Table 4 outlines the characteristics of the trial’s secondary publications in terms of types of analyses and their indicators of impact, stratified by authorship.

**Table 4:**
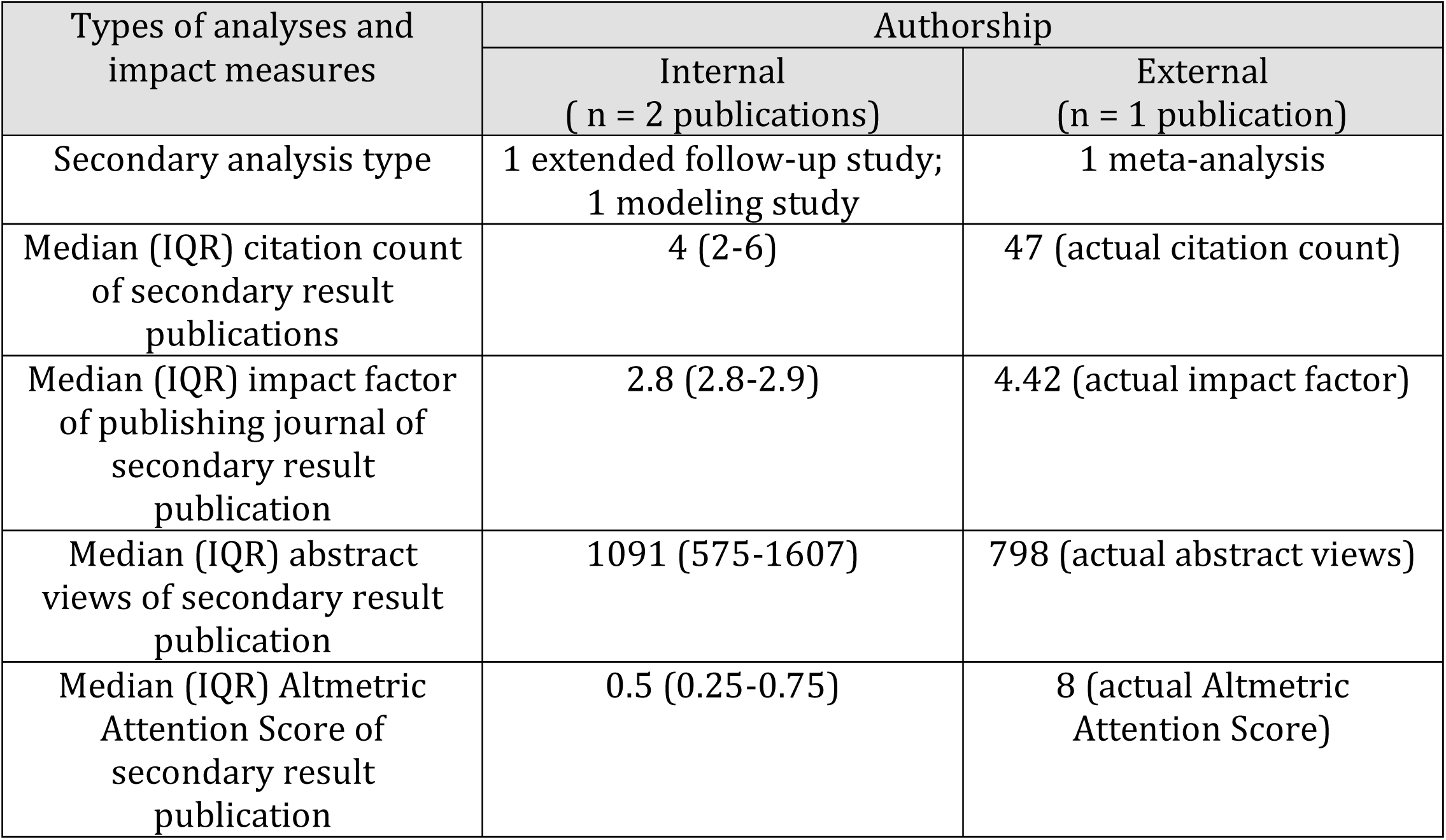
Types of analyses and impact measures of secondary publications authored by internal and external researchers for the trial Disease Management for Smokers in Rural Primary Care (NCI-funded)

| Types of analyses and impact measures | Authorship |  |
| --- | --- | --- |
|  | Internal<br>( n = 2 publications) | External<br>(n = 1 publication) |
| Secondary analysis type | 1 extended follow-up study;<br>1 modeling study | 1 meta-analysis |
| Median (IQR) citation count of secondary result publications | 4 (2-6) | 47 (actual citation count) |
| Median (IQR) impact factor of publishing journal of secondary result publication | 2.8 (2.8-2.9) | 4.42 (actual impact factor) |
| Median (IQR) abstract views of secondary result publication | 1091 (575-1607) | 798 (actual abstract views) |
| Median (IQR) Altmetric Attention Score of secondary result publication | 0.5 (0.25-0.75) | 8 (actual Altmetric Attention Score) |

The NCI-funded trial, *Enhancing Tobacco Use Treatment for African American Light Smokers*, had 13 total secondary publications, 12 of which were conducted by internal authors and 1 by external authors (Table 5). Of the 12 internal secondary publications, 9 reported secondary outcomes, 2 were pooled analyses and 1 was a sub-group analysis. The only shared data publication was a meta-analysis. The secondary internal publications had a median citation count of 20 (IQR, 8.5-27.5); median impact factor of 3.6 (IQR, 2.9-5.4); median abstract views of 21 (IQR, 2-313); and median Altmetric Attention Score of 1 (IQR, 0-3). The meta-analysis had a citation count of 37, was published in a journal with an Impact Factor of 4.1, had 9 abstract views and an Altmetric Attention Score of 34. Table 5 outlines the characteristics of the trial’s secondary publications in terms of types of analyses and their indicators of impact, stratified by authorship.

**Table 5:** Types of analyses and impact measures of secondary publications authored by internal and external researchers for the trial: Enhancing Tobacco Use Treatment for African American Light Smokers (NCI-funded)

| Types of analyses and impact measures | Authorship |  |
| --- | --- | --- |
|  | Internal<br>( n = 12 publications) | External<br>(n = 1 publication) |
| Secondary analysis type | 9 secondary outcome reporting; 2 pooled analyses; 1 sub-group analysis | 1 meta-analysis |
| Median (IQR) citation count of secondary result publications | 20 (8.5-27.5) | 37 (actual citation count) |
| Median (IQR) impact factor of publishing journal of secondary result publication | 3.6 (2.9-5.4) | 4.1 (actual impact factor) |
| Median (IQR) abstract views of secondary result publication | 21 (2-313) | 9 (actual abstract views) |
| Median (IQR) Altmetric Attention Score of secondary result publication | 1 (0-3) | 34 (actual Altmetric Attention Score) |

## Discussion

Among a matched sample of clinical trials completed in 2010 with total costs greater than or equal to $1,000,000 or total enrollment of 500 or more patients, we found that both models resulted in data sharing for a minority of eligible trials and generated few secondary data publications. Nevertheless, more shared data publications were associated with the NHLBI centralized model, suggesting that a centralized data sharing model may increase IPD accessibility and use.

The number of clinical trials sponsored by NHLBI and NCI that we identified for which data sharing would be required, either because the trial exceeded enrollment or cost thresholds, was smaller than we had anticipated. Due to the small sample size of matched trials in our study, we were precluded from making statistical comparisons and determining which data sharing model was more successful with respect to being associated with more shared data publications. For both models, approximately 21% of trials shared data, a rate that is quite low, although comparable with previous research.^22,23^ It is important to highlight this low rate of data shared from these large, resource-intensive trials that are publicly funded through the NIH, signifying their public health importance. Through personal communication with officials at BioLINCC, we confirmed that there were difficulties with initial implementation of the data sharing policy, particularly some challenges tracking trials that were eligible for sharing, which may explain the low proportion of trials sharing data via BioLINCC. Personal communication with authors of other NHLBI-funded trials that did not share their data at BioLINCC confirmed that 1 study was instead deposited data at dbGap, an NIH depository for genomic data.

As one would expect its centralized platform to be more accessible to the broader research community, it is encouraging to note that NCI’s data sharing model has evolved since Febuary 2017, into NCTN/NCORP Data Archive – a centralized repository where data can be requested, which is similar to BioLINCC’s data sharing model. It is expected that this shift to a centralized repository will increase the accessibility of these NCI-funded trials. The NCTN/NCORP Data Archive will initially house NCTN trials that were published on or after January 1, 2015.^20,24^

Our findings demonstrate that the opportunity for generating more scientific knowledge through clinical trial data sharing is yet to be maximized and the DOSE-AHF trial is an illustrative example highlighting the importance and value of sharing data from large clinical trials. A large number of shared data publications have thus far resulted from the the DOSE-AHF trial, research that would not have been possible without NHLBI and the study investigators sharing their data for external investigators to use for their own research studies. It is noteworthy that the Final NIH Policy for Data Management and Sharing, released on October 2020, effective from January 25, 2023, indicated that the NIH expects that researchers will “maximize appropriate data sharing” when developing their Data Management and Data Sharing Plans; an important step towards more open science.^12^

### Limitations

There are several limitations to our study. First, our study sample was limited to only 14 trials per NIH institute, precluding statistical comparison between the two groups and limiting the generalizability of our findings. However, this small sample size is partially attributable to our inclusion of only those trials expected to be of greatest importance to the clinical and research communities, as determined by trial costs and enrollment. Further, we would expect rates of results reporting, publication, and data sharing, and use of those shared data for secondary research, to be highest for these larger and more well-funded studies. As new NIH results reporting and data sharing policies are implemented, further research should examine if data are shared more readily and used more widely. Second, our assessments of publication and data sharing were conducted at a point in time. It is possibile that investigators later published or shared data from their trials. In fact, through personal communication with authors, investigators associated with one trial explained that the study was still ongoing and so had not yet been been published. Finally, the NCI data sharing model appeared to have been organized around Cooperative Groups, but not all NCI-funded trials included in our analysis were Cooperative Group trials. Nevertheless, all were still subject to the NIH data sharing policy.

## Conclusions

The true value in sharing data lies in the generation and dissemination of new knowledge resulting from shared data, not only in making data available to be shared. This study characterized the “outcomes” associated with two data sharing models used by the NHLBI and NCI, finding low rates of data sharing and numbers of shared data publications for the largest and most costly trials funded by the Intstitutes in 2010. This study was a direct response to the Institute of Medicine’s call for attention to this issue.^21^ However, while neither model resulted in the majority of trials being shared and both were associated with few secondary data publications, a larger number of shared data publications were associated with clinical trials, when shared, whose data were made available through the NHLBI centralized model. Future research is needed to further evaluate these and other existing data sharing models and initiatives to inform effective policies which maximize clinical trial data sharing, advancing the field’s understanding on how and to what extent IPD are being shared and used.

## Methods

### Study Sample – inclusion/exclusion

We assembled a sample of clinical trials sponsored by the NHLBI and NCI with a primary completion date between January 1, 2010 and December 31, 2010 and which had a project start date after May 1, 2006. We selected 2010 as the primary completion date year to ensure that sufficient time had passed for investigators to publish their primary findings, deposit and share their data, and for external authors to request these data and undertake and publish their secondary analyses. May 1, 2006 was selected as the earliest project start date, as the NHLBI data sharing policy was extended to grant-supported studies in 2005.

Eligible studies were identified using data downloaded from ClinicalTrials.gov and NIH RePORTER. We selected studies that for which NIH listed total costs greater than or equal to $1,000,000 in any given fiscal year, ensuring we identified those which had direct costs greater than or equal to $500,000 in any given fiscal year and with more than 500 participants. After a preliminary search, there were only two studies that fit both criteria; we revised our search to also include studies which fit either the sample size or cost criteria.

Once eligible studies were identified, we matched studies sponsored by each Institute based on either cost or size.

### Main Outcome Measures

#### Sharing of individual patient-level data and results reporting

For NHLBI-sponsored trials, we determined whether IPD were available for request on the BioLINCC website in March 2021. For trials not listed on BioLINCC, we contacted NHLBI staff to determine if the data were available for sharing. We checked NCI’s new data sharing portal, NCTN/NCORP Data Archive, for data from the NCI-sponsored studies.

We contacted corresponding authors (or secondary and senior authors if we could not locate the email address of the corresponding author) of all NHLBI-sponsored and NCI-sponsored studies for which we could not identify shared data in those public data sharing platforms, to confirm whether they had shared their data with other investigators. An initial email was sent, with a follow-up email if we did not hear back from author(s), in March 2021. No further follow-up emails were sent and we did not look for alternative email addresses when emails bounced back.

We determined whether results were reported on ClinicalTrials.gov for all trials in March 2021.

#### Publication Identification

##### Primary publication

We defined primary publications as those reporting the results for the primary outcome (i.e. the outcome used to determine sample size and study design). Primary publications were identified from the linked NIH RePORTER data. For trials without a primary publication listed, between September 2020 and March 2021, we searched ClinicalTrials.gov, MEDLINE, Google Scholar and a Google search first using the trial’s NCT number, then the title of the clinical trial. Trials without an identifiable primary publication were noted and corresponding authors (or secondary and senior authors if we could not locate the email address of the corresponding author) were contacted to request a copy of the trial’s primary publication. An initial email was sent, with a follow-up email if author(s) did not respond to our request, in March 2021. No further follow-up emails were sent and we did not look for alternative email addresses when email(s) bounced back.

##### Secondary publications

We defined secondary publications as those reporting IPD analyses that were separate from that reported in the primary publication. To identify secondary publications, between September 2020 and March 2021, we searched MEDLINE for all publications citing the primary publication and/or listing the trial’s NCT identifier, manually screening the titles and abstracts to flag eligible studies. Full-text articles of flagged citations were reviewed for eligibility. We excluded publications reporting the results of pilot studies, feasibility studies, study protocols, interim analyses, and non-human studies. We defined shared data publications as those secondary publications reporting IPD analyses that were separate from that reported in the primary publication, authored only by external authors. Even if only one author from the primary publication was included on the secondary publication, we categorized the study as a secondary publication, not a shared data publication.

#### Impact

For each primary publication, we extracted the authorship, publishing journal, and intervention type (i.e., interventional or observational). For each secondary publication, we extracted the type of analysis (i.e. extended follow-up, reanalysis of primary outcome, secondary outcome reporting, subgroup analysis, pooled analysis, predictive analysis, meta-analyses); authorship; publishing journal; and acknowledgement of IPD source, times cited, impact factor of the publishing journal, abstract views, and the Altmetric Attention Score, an indicator of the amount of attention the articles have received.

Two investigators (ARF, ADZ) identified eligible trials. One investigator (ARF) identified publications and performed data abstraction, which was then verified by another investigator (AE) to ensure accuracy. Throughout the data collection process, a random sample of abstracted data was periodically examined by a senior author (JSR) to assess accuracy.

### Statistical Methods

We used descriptive statistics to characterize the number of trials that published their findings, reported their results, shared their IPD, and had secondary publications resulting from shared data (external or internal). We also calculated the median and interquartile range (IQR) for the time from primary study completion to primary publication for those trials that published their findings. For those trials that had secondary publications with external authors, we characterized the type of studies conducted and calculated the median and IQR for number of secondary publications, citation counts, impact factor of publishing journal, abstract views and Altmetric Attention Score. All analyses were conducted using Microsoft Excel, Version 16.46.

## Data Availability

The data generated from this study are available at: https://osf.io/t8b6g/?view_only=e08e0fb3c4254343b610beaf10027b52 (view-only link as data will be made publicly available once accepted for publication).

## Code sharing statement

There is no custom computer code used to analyze the data generated from this study; all analyses were conducted using Microsoft Excel, Version 16.46.

